# Maternal body mass index in early pregnancy and offspring risk of multiple sclerosis: A population-based cohort study

**DOI:** 10.64898/2026.10.15.26365617

**Authors:** Huiling Xu, Neda Razaz, Katarina Remaeus, Jimin J Lee, Katharina Fink, Kyla A. McKay

## Abstract

**What is already known on this topic:**

- Prenatal and perinatal factors have been linked to offspring MS risk, but evidence has focused mainly on maternal complications or birth outcomes.
- Maternal BMI has been associated with other immune-mediated diseases in offspring, but large-scale evidence linking early-pregnancy BMI to offspring MS risk is lacking.

**What this study adds:**

- In a nationwide cohort, higher maternal BMI in early pregnancy was associated with increased offspring MS risk.
- This association persisted after excluding pregnancies with major complications, suggesting it is not fully explained by adverse pregnancy outcomes.
- These findings provide the first large-scale evidence for the possibility that susceptibility to MS may begin to be shaped in early gestation.

**Objective:** To examine whether prospectively recorded maternal BMI in early pregnancy is associated with offspring multiple sclerosis (MS) risk, including childhood-onset and adult-onset MS.

**Design:** Nationwide population-based cohort study.

**Setting:** Swedish national health and demographic registers.

**Participants:** 1,524,863 singleton children born in Sweden from 1992 to 2007 and followed through 2025.

**Exposures:** Maternal BMI recorded at the first antenatal visit, categorised as underweight (<18.5), normal weight (18.5-24.9), overweight (25.0-29.9), and obesity (≥30.0), and also modelled continuously.

**Main outcome measures:** MS in offspring, identified as at least three ICD-10 G35 codes recorded on separate dates. Cox models estimated adjusted hazard ratios (HR) and 95% confidence intervals (CI) adjusting for confounders.

**Results:** During follow-up, 1539 offspring (0.1%) were diagnosed with MS at a median age of 24 years (interquartile range, 20-27 years). MS incidence rates by maternal BMI category were 0.37 per 10,000 person-years for both underweight, and normal weight, 0.42 for overweight, and 0.45 for obesity. Compared with normal weight women, maternal overweight and obesity were associated with higher hazards of offspring MS, with adjusted HRs of 1.20 (95% CI, 1.05 to 1.37) and 1.35 (95% CI, 1.11 to 1.64), respectively. Onset-specific analyses showed similar findings for adult-onset MS and less precise but directionally consistent estimates for paediatric-onset MS. Each 1-unit increase in maternal BMI was associated with a 3%, 4%, and 3% higher hazard of overall, paediatric-onset, and adult-onset offspring MS, respectively.

**Conclusions:** In this population-based cohort study, higher maternal BMI were associated with an increased rate of MS in offspring. The findings provide large-scale prospective evidence that maternal adiposity may be related to offspring MS risk and support further investigation of the prenatal metabolic environment in the early-life origins of MS.

## Introduction

Multiple sclerosis (MS) is a complex immune-mediated disorder of the central nervous system (CNS), characterized by inflammation, demyelination, and neurodegeneration.^1^ Globally, MS affects an estimated 2.8 million people with onset typically occurring between 18 and 40 years of age.^2^ As the disease progresses, MS frequently results in impairments in mobility and cognition, adversely affecting quality of life and daily functioning.^3^

Recent evidence suggests that susceptibility to MS may originate during the prenatal period.^4^ Large nationwide studies have reported that maternal diabetes, large or small size for gestational age, and mode of delivery may be associated with MS risk in offspring.^4–6^ Each of these factors is associated with maternal body mass index (BMI) and may represent downstream consequences of maternal BMI.^7^ ^8^ Overweight and obesity are increasingly common in women.^9^ High maternal BMI has been linked to adverse health outcomes in offspring, including in other immune-mediated diseases in offspring, such as type 1 diabetes and childhood asthma.^10–12^ Taken together, these findings suggest that maternal BMI may be associated with MS risk in offspring. Although maternal BMI may play an important role in this process, its potential contribution remains largely unexplored. Only two previous studies, both based on retrospectively collected maternal BMI information, have reported inconsistent findings.^13^ ^14^ To date, no nationwide study using prospectively collected BMI data has been conducted.

Therefore, our study aimed to investigate the association between early pregnancy BMI and the rate of MS in offspring, using data from a nationwide registry-based cohort including all singleton births in Sweden between 1992 and 2007, with follow-up through 2025.

## Method

### Study design and data source

This nationwide population-based cohort study accessed the following registers: the Medical Birth Register,^15^ National Patient Register,^16^ Total Population Register,^17^ the Swedish Cause of Death Register,^18^ and Longitudinal Integrated Database for Health Insurance and Labor Market Studies (LISA).^19^ All registers were linked through the unique personal identification number assigned to each Swedish resident. The study was approved by the Swedish Ethical Review Authority. The requirement for informed consent was waived as only anonymized registry data were used.

### Study population

Women with live singleton children born between 1992 and 2007 were included. Birth years from 1992 to 2007 were selected because maternal weight and self-reported height at the first antenatal visit have been recorded since 1992,^20^ and this range ensured that all offspring had reached 18 years of age by the end of follow-up in 2025. Offspring with no observable follow-up time were excluded (Supplementary Figure 1). The study followed the Strengthening the Reporting of Observational Studies in Epidemiology (STROBE) reporting guidelines.^21^

### Exposure

Early pregnancy BMI was calculated as weight (kilogram) measured at the first prenatal visit (in light clothing) divided by the square of self-reported height (meters) obtained from the same visit. In this study, the first prenatal visits took place at a median gestational age of 10 weeks, with an interquartile range of 9-12 weeks. Given the high correlation of self-reported height across pregnancies within the same mother,^22^ the median height across pregnancies was used for multiparous women to reduce measurement error.^23^ Early pregnancy BMI was categorized into underweight (<18.5 kg/m^2^), normal weight (18.5 to 24.9 kg/m^2^; reference), overweight (25.0 to 29.9 kg/m^2^), and obese (≥30.0 kg/m^2^), according to WHO BMI classification.^24^ Early pregnancy BMI was also considered as a continuous variable, to explore a potential non-linear relationship using restricted cubic splines.

### Covariates

Maternal age at delivery was calculated by subtracting the mother’s date of birth from the date of delivery. Information on cohabitation with the child’s father, an indicator of socioeconomic status,^25^ was collected at the first prenatal visit. Mothers who reported smoking either at the first prenatal visit or at 30-32 weeks of gestation were classified as smokers, whereas those reporting no smoking were classified as non-smokers.^26^ Parity was defined as the total number of births per mother and categorized as nulliparous or multiparous. Mothers’ region of birth (categorized as Nordic-born including Sweden, Denmark, Norway, Finland, Iceland and non-Nordic-born) and educational level (categorized as ≤12 years and >12 years [equivalent to university/college]) were also considered. Maternal MS was defined using the same criteria as for children (described below). Infant sex and year of birth were obtained from delivery care records.

Gestational age was categorized as term (≥37 weeks) or preterm (<37 weeks). In Sweden, all pregnant women are offered ultrasound-based pregnancy dating no later than the early second trimester, with an acceptance rate of approximately 95%.^27^ Gestational age in completed weeks was primarily determined by early second-trimester ultrasound (87.7%), followed by the date of the last menstrual period (7.4%) and postnatal assessment (4.9%).

In addition to the covariates included in the main models, several maternal and neonatal complications were reported to further characterize the study population. These variables were included for descriptive purposes only and were not included in the adjusted models because they were not, by definition, confounders. They included diabetes (gestational and mellitus), hypertension (pregestational and preeclampsia), mode of delivery, macrosomia (birth weight ≥4000 g), birth weight for gestational age according to sex-specific Swedish reference curves for normal fetal growth,^28^ any neonatal infections within 28 days of birth, low Apgar score at 5 minutes (<7). ICD codes for these maternal and neonatal complications are provided in Supplementary Table 1 in the Supplement. The categorization of covariates is presented in Tables 1 and 2.

**Table 1.** The distribution of maternal characteristics according to offspring MS status.

| Maternal characteristics | Offspring with MS | Offspring without MS |
| --- | --- | --- |
| **Total, n (%)** | 1539 (0.1) | 1523324 (99.9) |
| **Maternal age, y** |  |  |
| Mean (SD) | 28.8 (5.2) | 29.4 (5.1) |
| ≤19 | 42 (2.7) | 29635 (1.9) |
| 20-24 | 295 (19.2) | 233816 (15.3) |
| 25-29 | 533 (34.6) | 519391 (34.1) |
| 30-34 | 436 (28.3) | 490763 (32.2) |
| ≥35 | 233 (15.1) | 249719 (16.4) |
| **Region of birth** |  |  |
| Nordic | 1207 (78.4) | 1293838 (84.9) |
| Non-nordic | 331 (21.5) | 228654 (15.0) |
| Missing | <5 (0.1) | 832 (0.1) |
| **Education, y** |  |  |
| >12 | 617 (40.1) | 744897 (48.9) |
| ≤12 | 918 (59.6) | 773407 (50.8) |
| Missing | <5 (0.3) | 5020 (0.3) |
| **Mother cohabits with partner** |  |  |
| Yes | 1364 (88.6) | 1349010 (88.6) |
| No | 83 (5.4) | 74420 (4.9) |
| Missing | 92 (6.0) | 99894 (6.6) |
| **Parity** |  |  |
| Nulliparous | 605 (39.3) | 648838 (42.6) |
| Multiparous | 934 (60.7) | 874486 (57.4) |
| **Maternal height, cm** |  |  |
| Mean (SD) | 165.8 (6.4) | 166.3 (6.3) |
| ≤159cm | 234 (15.2) | 186175 (12.2) |
| 160-164 | 397 (25.8) | 375860 (24.7) |
| 165-169 | 404 (26.3) | 438266 (28.8) |
| ≥170 | 437 (28.4) | 469852 (30.8) |
| Missing | 67 (4.4) | 53171 (3.5) |
| **Maternal MS** |  |  |
| No | 1527 (99.2) | 1522268 (99.9) |
| Yes | 12 (0.8) | 1043 (0.1) |
| Missing | 0 (0.0) | 13 (0.0) |
| **Smoking during pregnancy** |  |  |
| No | 1203 (78.2) | 1244184 (81.7) |
| Yes | 274 (17.8) | 202734 (13.3) |
| Missing | 62 (4.0) | 76406 (5.0) |
| **Maternal Infection** |  |  |
| No | 1474 (95.8) | 1460059 (95.8) |
| Yes | 65 (4.2) | 63265 (4.2) |
| **Diabetes disease** |  |  |
| No | 1517 (98.6) | 1503313 (98.7) |
| Pregestational diabetes | 6 (0.4) | 6436 (0.4) |
| Gestational diabetes | 16 (1.0) | 13575 (0.9) |
| **Hypertensive disorders** |  |  |
| No | 1485 (96.5) | 1471444 (96.6) |
| Pregestational hypertension | 10 (0.6) | 9258 (0.6) |
| Preeclampsia | 44 (2.9) | 42622 (2.8) |

**Table 2.**
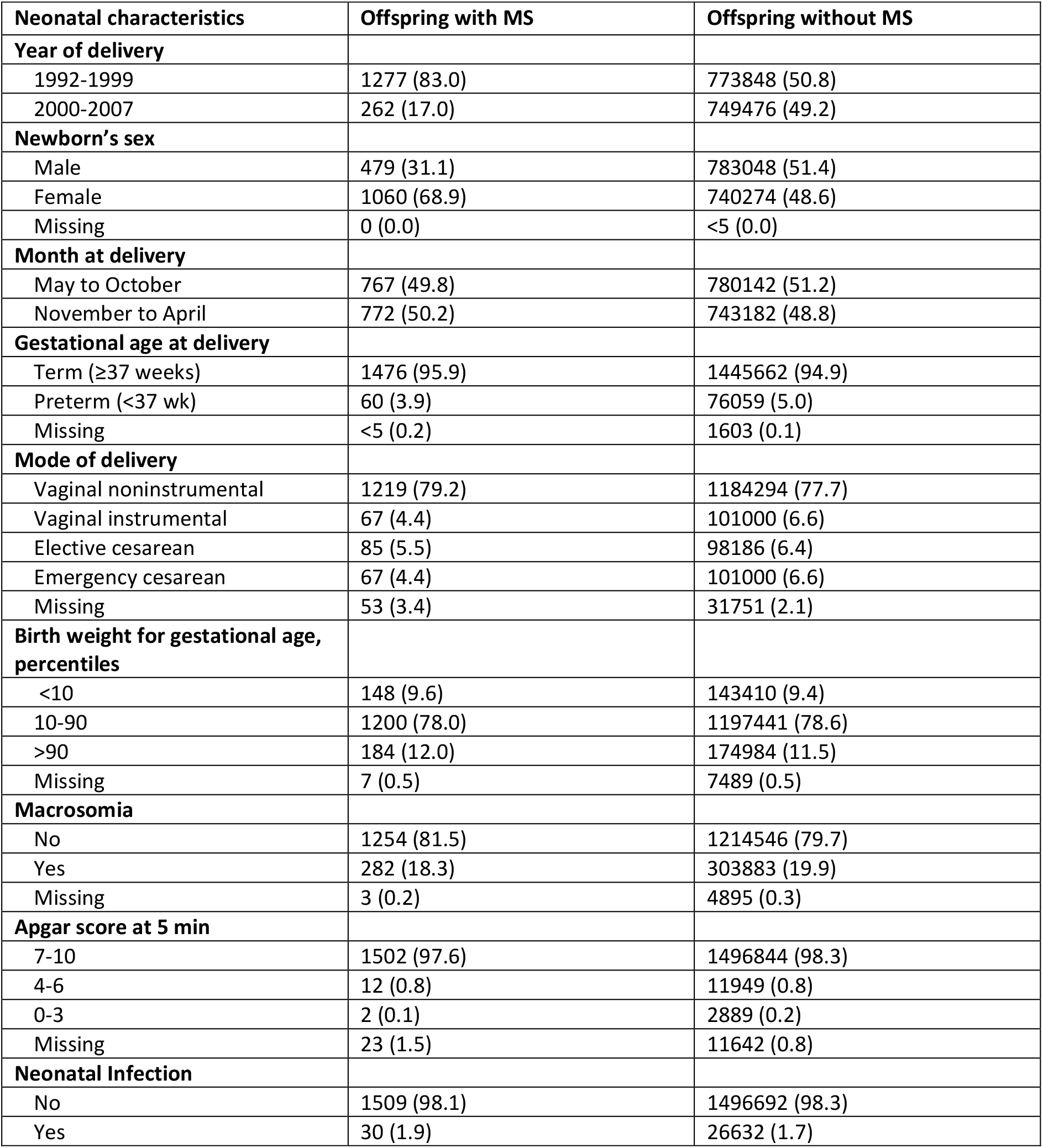
Delivery and neonatal characteristics according to offspring MS status.

### Outcome

MS cases (and maternal MS) were identified from the National Patient Register using ICD-9 (340) and ICD-10 (G35) codes, including both primary and secondary diagnoses. A validated case definition was applied, requiring at least three MS-coded visits recorded on separate dates (positive predictive value: 98.8).^29^ Among individuals fulfilling this definition, the index date was defined as the date of the first recorded central nervous system demyelinating event (ICD-9: 377D, 323X, 341W, 341X, 341A; ICD-10: H46, H47,H48, G373, G369,G378, G36, G360). MS cases were further classified as paediatric-onset MS (POMS) and adult-onset MS (AOMS), defined as first CNS demyelinating code before 18 years of age, and at 18 years of age or older, respectively.

### Statistical analysis

Baseline maternal characteristics, pregnancy complications, and neonatal complications were described by MS separately. Baseline covariates were also described across maternal BMI categories. Continuous variables are presented as means with standard deviations (SD) or medians with interquartile ranges (IQR), and categorical variables as counts with percentages.

We estimated person-time incidence rates of MS. In the overall analysis, offspring were followed from birth until the first recorded diagnosis of MS, emigration, death, or 31 December 2025 (end of data availability), whichever occurred first. For the pediatric-onset MS analysis, offspring were followed from birth until the first recorded diagnosis of MS, emigration, death, 18 years of age, or December 31, 2025, whichever occurred first. For the adult-onset MS analysis, offspring who were alive, residing in Sweden, and free of MS at 18 years of age entered follow-up at 18 years of age and were followed until the first recorded diagnosis of MS, emigration, death, or December 31, 2025, whichever occurred first. Those with no observable follow-up time after age 18 years were excluded from the adult analysis dataset. The association between maternal BMI and the rate of MS in offspring was examined using Cox proportional hazards models. The proportional hazards assumption was assessed using Schoenfeld residuals. No evidence of violation was observed for the exposure. All estimates were presented as hazard ratios (HRs) with 95% confidence intervals (CIs). Adjusted HRs were obtained from multivariable Cox models with covariates including maternal age, parity, mother’s region of birth, education level, cohabitation with partner, height, smoking status, maternal MS, and year of delivery. Given that the analyses were performed at the individual pregnancy level, robust standard errors clustered by maternal serial number were used to address potential correlations between repeated pregnancies within the same mother. Maternal BMI was primarily analyzed as a categorical variable. To explore potential non-linear associations, maternal BMI was additionally modelled as a continuous variable using restricted cubic splines with four knots placed at the 5th, 35th, 65th, and 95th percentiles of the BMI distribution. Overall association and non-linearity were assessed using Wald tests. In the absence of evidence for a non-linear association, the HRs per 1-unit increase in maternal BMI was estimated by treating BMI as a continuous variable.

Given that gestational age is related to immune system development,^30^ ^31^ and maternal BMI is associated with preterm delivery,^32^ we repeated the primary analyses restricted to term births (≥37 weeks) to assess whether the association between maternal BMI and offspring MS persisted independent of preterm-related factors. Separate analyses among preterm births were not performed because of the limited number of cases. In addition, we further restricted the analyses to offspring born at term (≥37 weeks), neither large or small for gestational age (LGA or SGA, respectively), and without maternal diabetes, hypertension or preeclampsia to assess whether the association persisted in this low-risk subgroup. We also performed separate analyses among nulliparous and multiparous women to assess whether the association between maternal BMI and offspring MS varied by parity, given potential differences in maternal BMI and pregnancy characteristics between nulliparous and multiparous women.^33^ Furthermore, we performed sex-stratified analyses to assess whether the association between maternal BMI and offspring MS differed between male and female offspring. Approximately 15% of pregnancies did not have early-pregnancy BMI data. To address the potential impact of missing data on the estimated association, we performed multiple imputation as a sensitivity analysis. Missing values for BMI and covariates were imputed using multiple imputation by chained equations before fitting the multivariable Cox regression models.34 The imputation model included maternal age, country of origin, education level, cohabitation with a partner, parity, height, weight measured at the first antenatal care visit and before delivery, smoking during pregnancy, year of delivery, infant weight, infant length, gestational length, offspring MS diagnosis, and the Nelson-Aalen estimate of the cumulative baseline hazard.35 Multiple imputation was performed with 10 imputed datasets, and estimates from the multivariable models were pooled using Rubin’s rules as implemented in the mice package in R.

All analyses were performed using R version 4.5.3.

### Patient and public involvement

No patients or members of the public were directly involved in the design, conduct, analysis, reporting, or dissemination plans of this registry-based study. The study used pseudonymised population register data, which precluded direct involvement of the individuals included in the data.

## Results

Among 1,524,863 singleton children born in Sweden between 1992 and 2007, 1539 developed MS by December 31, 2025, including 1060 female and 479 male cases. Among all MS cases, the median age at diagnosis was 23.5 years (IQR, 20.0-26.8 years). Of all MS cases, 220 were classified as POMS, and 1315 as AOMS. Among POMS cases, the median age at the index date was 16.4 years (IQR, 14.7-17.2 years); 148 cases were female and 72 were male. Among AOMS cases, the median age at the index date was 24.5 years (IQR, 21.8-27.3 years), and 909 cases were female and 406 were male.

**Tables 1 and 2** present the distribution of maternal, delivery, and neonatal characteristics according to offspring MS status. Mothers of offspring with MS were more likely to be younger at delivery, have a lower education level (≤12 years), multiparity, shorter stature, a history of MS, and to smoke during pregnancy. Offspring with MS were also more likely to be female, born between November and April, and delivered vaginally without instrumental assistance. The distribution of maternal characteristics across maternal BMI categories are listed in Supplementary Table 2 in the supplementary.

Among women with available early pregnancy BMI data, the mean early pregnancy BMI was 24.2 (SD, 4.2), and the proportions of BMI categories were 2.6% underweight (n=34076), 64.2% normal weight (n=829967), 23.8% overweight (n = 307748), and 9.4% obesity (n=121014).

In the overall MS analysis, the numbers of MS cases across the underweight, normal-weight, overweight, and obesity categories were 32, 776, 320, and 132, respectively, and the corresponding incidence rates per 10000 person-years were 0.37, 0.37, 0.42, and 0.45. Compared with normal BMI, maternal overweight and obesity were associated with higher hazards of offspring MS, with adjusted HRs of 1.20 (95% CI, 1.05-1.37) and 1.35 (95% CI, 1.11-1.64), respectively, whereas no clear association was observed for underweight. In the adult-onset analysis, similar associations were observed. For paediatric-onset analysis, the point estimates suggested a positive association for overweight and obesity; however, the estimates were imprecise, with wide confidence intervals, and did not reach statistical significance, likely owing to the smaller number of events (**Figure 1**).

**Figure 1.**
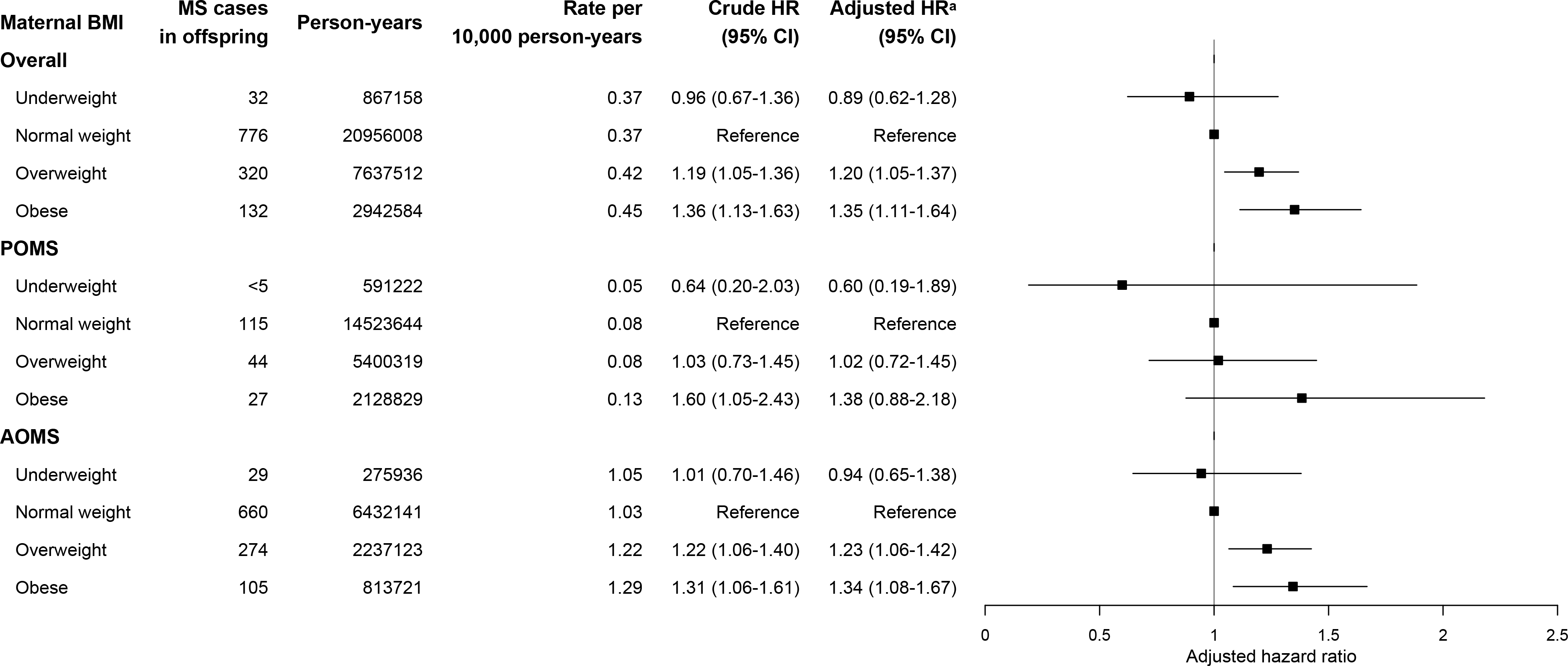
Early pregnancy BMI and rate of MS in singleton offspring born 1992-2007 in Sweden. a. Adjusted for maternal age, parity, mother’s region of birth, education level, cohabitation with partner, height, smoking status, maternal MS, and year of delivery.

When maternal BMI was modelled using spline terms, there was no evidence of a non-linear association with overall MS (P for non-linearity = 0.30), POMS (P for non-linearity = 0.79), or AOMS (P for non-linearity = 0.16). The spline curves suggested a generally increasing association between maternal BMI and the hazard of overall MS, POMS, and AOMS (Figure 2). For POMS, this positive pattern appeared more evident in the spline analysis than in the categorical analysis; however, at the extreme high end of maternal BMI, the confidence intervals were wide and the number of events was limited, indicating that the estimates in this range were imprecise and should be interpreted cautiously despite the apparent upward trend. In the adjusted model, each 1-unit increase in maternal BMI was associated with a 3% higher hazard of overall MS, a 4% higher hazard of POMS, and a 3% higher hazard of AOMS (overall MS: aHR, 1.03; 95% CI, 1.02-1.04; POMS: aHR, 1.04; 95% CI, 1.01-1.07; AOMS: aHR, 1.03; 95% CI, 1.01-1.04).

**Figure 2.**
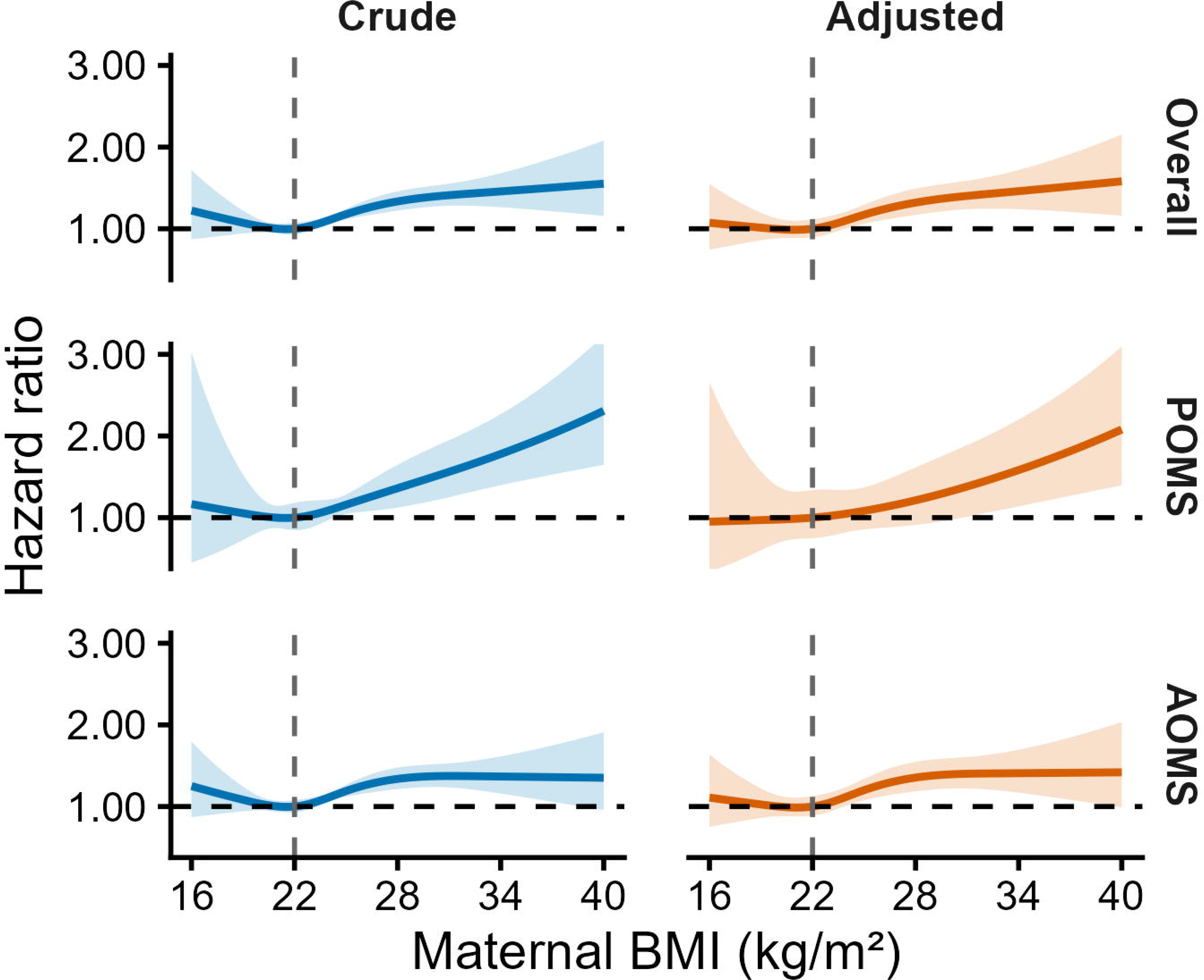
Non-linear association between maternal BMI and rate of MS in offspring. Hazard ratios (HRs) for multiple sclerosis (MS) according to maternal BMI modelled as a continuous variable using restricted cubic splines, with BMI 22 kg/m² as the reference. Shaded areas indicate 95% CIs. Adjusted for maternal age, parity, maternal region of birth, education level, cohabitation with a partner, height, smoking status, maternal MS, and year of delivery.

When analyses were restricted to term births, the estimates for maternal obesity, compared with normal weight, were slightly higher than observed in the overall cohort (aHR for overall MS, 1.40; 95% CI, 1.15-1.71; aHR for POMS, 1.47; 95% CI, 0.93-2.33; aHR for AOMS, 1.39; 95% CI, 1.11-1.73) (**Figure 3**). Similar patterns were observed when analyses were further restricted to term appropriate-for-gestational-age births uncomplicated by maternal diabetes, hypertension, or preeclampsia (aHR for overall MS, 1.43; 95% CI, 1.13-1.82; aHR for POMS, 1.32; 95% CI, 0.74-2.35; aHR for AOMS, 1.46; 95% CI, 1.13-1.90) (Supplementary Figure 2).

**Figure 3.**
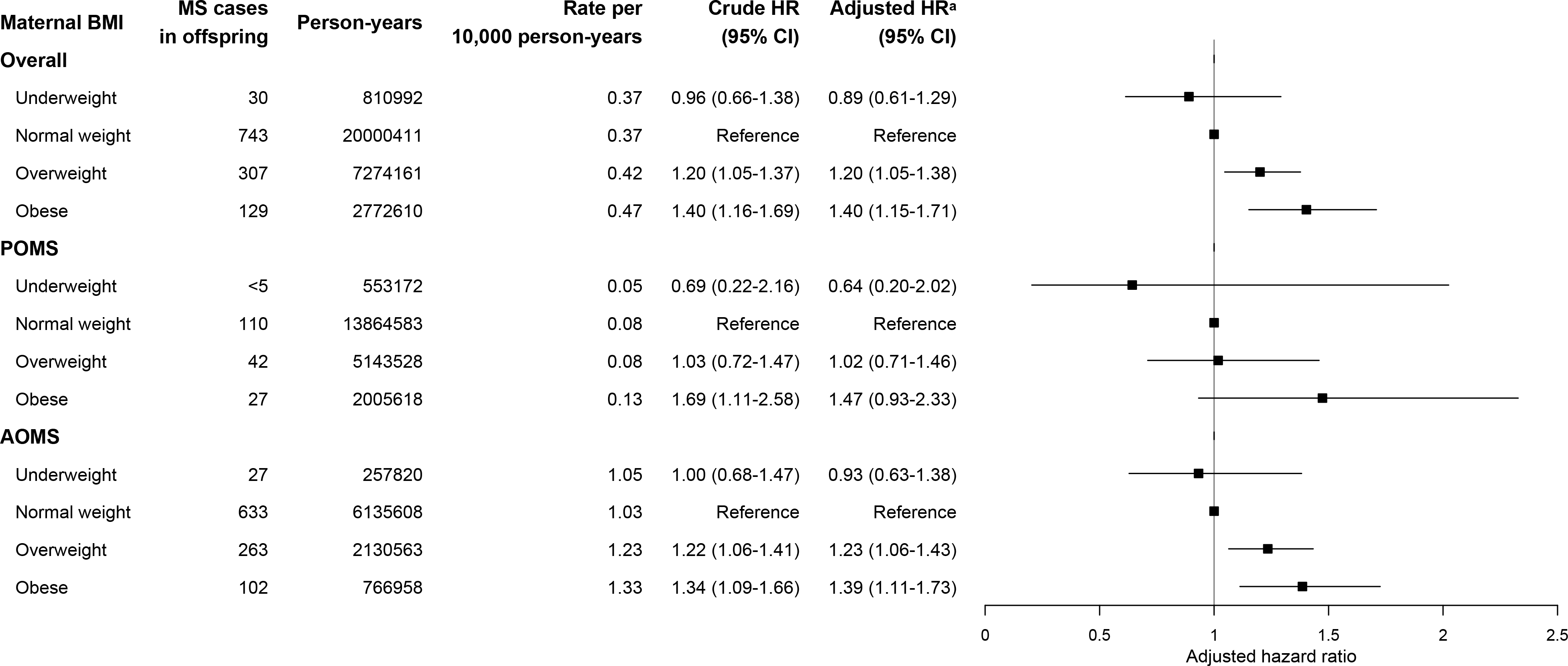
Early-pregnancy BMI and rate of MS in term-born singleton offspring in Sweden, 1992-2007. a. Adjusted for maternal age, parity, mother’s region of birth, education level, cohabitation with partner, height, smoking status, maternal MS, and year of delivery.

In analyses stratified by parity, the direction and overall pattern of associations were similar to those in the primary cohort, although estimates were imprecise and did not reach statistical significance among nulliparous women, likely due to the smaller number of events (Supplementary Figure 3). Similarly, in analyses stratified by offspring sex, the direction and overall pattern of associations were comparable to those in the primary cohort. Estimates among women with male offspring were imprecise and not statistically significant, likely owing to the smaller number of MS cases in men, whereas the effect size appeared larger among women with female offspring (Supplementary Figure 4). Results after multiple imputation were materially unchanged, with slightly higher incidence rates per 10000 person-years (Supplementary Table 3).

## Discussion

### Statement of principal findings

In this nationwide Swedish cohort study, maternal overweight and obesity were associated with higher rates of overall MS in offspring, particularly among those born at term. Similar associations were observed for adult-onset MS, while estimates for paediatric-onset MS showed a broadly similar pattern but were less precise and were statistically non-significant, likely because of smaller number of events. Complementing these findings, spline analyses suggested a generally increasing and approximately linear association between maternal BMI and MS risk. Each 1-unit increase in maternal BMI was associated with a 3%, 4%, and 3% higher hazard of overall MS, POMS, and AOMS, respectively.

### Strengths and weaknesses of the study

The study has several strengths. The nationwide cohort included over 1.5 million births identified through the Swedish Medical Birth Register. Unlike all previous studies of this relationship, maternal information was collected prospectively during pregnancy, and maternal weight was measured in early pregnancy rather than self-reported, thereby eliminating recall bias. Linkage with other national registers provided detailed data on mothers and offspring, enabling adjustment for a wide range of potential confounders. In addition, the publicly funded antenatal care system ensures near-complete population coverage, with over 99% of pregnancies captured in the Medical Birth Register, minimising selection bias.

Several limitations should be considered when interpreting the findings. Approximately 15% of women had missing information on BMI. If missingness was related to both maternal BMI and offspring MS risk, complete-case analysis could yield biased estimates. To address this, multiple imputation was performed and the results were materially unchanged, suggesting that missing data did not substantially influence our findings. In addition, the small number of pediatric-onset MS cases resulted in limited precision, and the estimates for this group should be interpreted as hypothesis-generating and warrant further investigation in larger studies. Another limitation is that follow-up extended only into young adulthood due to BMI data availability, meaning that the eldest a person could be at MS recognition was 33 years of age, which represents the mean age at MS onset. Accordingly, our findings primarily reflect MS diagnosed in adolescence and early adulthood and may not be generalizable to MS diagnosed later in adulthood. Early-pregnancy BMI was used as a proxy for pre-pregnancy BMI and may slightly overestimate true pre-pregnancy BMI, although weight change during early pregnancy is generally minimal.^36^ We lacked information on offspring adiposity during childhood and adolescence and therefore could not assess whether later offspring obesity mediated the association between maternal early-pregnancy BMI and offspring MS risk. In addition, residual confounding by shared familial and lifestyle factors cannot be excluded. Finally, our method of identifying MS was intended to capture all cases, however, there remains a risk of misidentification. To address this concern, we used a stringent case definition of MS with a high positive predictive value. Finally, as Sweden has a universal healthcare system, our findings may not be directly generalizable to populations with different healthcare systems.

### Strengths and weaknesses in relation to other studies

Evidence on the association between early-pregnancy BMI and offspring MS risk remains limited and inconsistent; only two studies have examined this relationship, both relying upon maternal recall of their weight during pregnancy. A US-based study found no association between maternal pre-pregnancy BMI and pediatric-onset MS.^14^ However, the mothers were questioned on their weight at pregnancy, on average, 15 years post-pregnancy. Another US-based study similarly relied on self-reported questionnaire data and found that maternal underweight was associated with a decreased rate of adult-onset MS, whereas maternal overweight/obesity was associated with an increased rate, but event rates for both were small (7 MS cases born to underweight and 20 born to overweight/obesity mothers).^13^ Compared with these studies, the present study used prospectively collected antenatal data and measured maternal weight in early pregnancy, reducing recall bias and improving exposure assessment.

Our findings are also consistent with emerging register-based evidence that intrauterine metabolic factors may be related to later MS risk. A recent study from Norway with a comparable data structure to the present study found that maternal diabetes and large-for-gestational-age births were associated with increased rates of adult-onset MS in offspring, whereas small-for-gestational-age births were associated with decreased rates of adult-onset MS. Maternal BMI is associated with maternal diabetes and offspring birth weight^37–39^ and may therefore represent an upstream determinant of an adverse intrauterine metabolic environment. Our findings extend this line of research by suggesting that maternal BMI itself is associated with the risk of MS in offspring.

### Meaning of the study

These findings should not be interpreted as evidence that reducing maternal BMI in early pregnancy would prevent MS in offspring, as establishing causality would require studies specifically designed to assess intervention effects. Rather, using prospectively collected BMI data from national registries, this study provides the first large-scale evidence linking early-pregnancy maternal overweight and obesity to offspring MS risk. This is important because previous studies of prenatal and perinatal factors in relation to offspring MS have largely focused on maternal complications or birth outcomes, such as maternal diabetes or fetal growth.^4–6^ By contrast, our findings shift attention to maternal adiposity in early pregnancy as a more upstream exposure that may precede these later pregnancy and birth-related factors.

The persistence of the association after restricting the sample to term pregnancies without complications suggests that the observed association is unlikely to be fully explained by adverse pregnancy outcomes alone. This raises the possibility that susceptibility to MS may begin to be shaped earlier in gestation, with early pregnancy representing a potentially important window for fetal immune programming.

The mechanisms underlying this association remain uncertain, but several pathways are biologically plausible. Maternal obesity induces a proinflammatory intrauterine environment that may alter fetal immune programming through placental inflammation, epigenetic modification, and impaired intestinal barrier development.^40–42^ In addition, maternal obesity is associated with an increased risk of obesity in the offspring, which itself has been linked to MS risk.^43^ Finally, higher BMI is associated with lower circulating vitamin D levels,^44^ which may diminish the potential protective role of vitamin D against MS.^45^

Notably, our findings are consistent with a broader pattern linking maternal obesity to immune-mediated diseases in offspring, including type 1 diabetes and childhood asthma, with comparable effect sizes,^12^ ^46^ suggesting shared pathways through which maternal adiposity may program offspring immune system development rather than a disease-specific mechanism.

### Unanswered questions and future research

This study provides new evidence linking maternal adiposity in early pregnancy to offspring MS risk, while raising important questions about causality, underlying mechanisms, and the timing of MS susceptibility. Because MS is a rare outcome, particularly when examined by age at onset or within pregnancy subgroups, larger datasets, longer follow-up, and international collaboration between population-based registers will be needed. Such efforts could provide sufficient statistical power to clarify the association with paediatric-onset MS, assess MS diagnosed later in adulthood, and better address residual genetic, familial, and lifestyle confounding. In addition, linking population-based registers with antenatal laboratory, placental, and offspring health data will be needed to investigate potential mediating pathways, including maternal metabolic and inflammatory markers, vitamin D status, placental function, offspring adiposity, and markers of offspring immune development. Such work may help determine whether early pregnancy represents a critical window in the development of MS susceptibility.

## Conclusion

In this nationwide population-based cohort study, higher maternal BMI in early pregnancy was associated with an increased rate of MS in the offspring. While residual confounding cannot be excluded, these findings add to growing evidence linking the prenatal metabolic

environment to offspring immune-mediated disease and may inform future preventive strategies.

## Supporting information

Supplementary Table 1. ICD-9 and ICD-10 codes for maternal and neonatal diseases Supplementary Table 2. Distribution of maternal characteristics acros

## Data Availability

The data are not publicly available due to privacy and data protection regulations and cannot be shared by the authors.

## Author Contribution statements (rank by first name’s alphabetical order)

Designed the study; Huiling Xu and Kyla McKay.

Analyzed data or performed statistical analysis; Huiling Xu.

Drafting of the manuscript; Huiling Xu.

Critical revision of the manuscript for important intellectual content; all authors

Huiling Xu and Kyla McKay are the guarantors of this work and, as such, had full access to all the data in the study and take responsibility for the integrity of the data and the accuracy of the data analysis.

