## Supplementary Table 1. ICD-9 and ICD-10 codes for maternal and neonatal diseases Supplementary Table 2. Distribution of maternal characteristics acros for "Maternal body mass index in early pregnancy and offspring risk of multiple sclerosis: A population-based cohort study"

**Supplementary Online Content**
Huiling Xu; Neda Razaz; Katarina Remaeus; Jimin J Lee; Katharina Fink; Kyla A. McKay

**Supplementary Table 1.** ICD-9 and ICD-10 codes for maternal and neonatal diseases

**Supplementary Table 2.** Distribution of maternal characteristics across early-pregnancy body mass index categories

**Supplementary Table 3.** Early-pregnancy body mass index and rates of multiple sclerosis in the overall cohort after multiple imputation

**Supplementary Figure 1.** Flowchart of inclusion criteria and final study population

**Supplementary Figure 2.** Early-pregnancy body mass index and rates of multiple sclerosis in term appropriate-for-gestational-age births uncomplicated by maternal diabetes, hypertension, or preeclampsia

**Supplementary Figure 3.** Early-pregnancy body mass index and rates of multiple sclerosis in offspring among nulliparous and multiparous women

**Supplementary Figure 4.** Early-pregnancy body mass index and rates of multiple sclerosis in male versus female offspring.

Supplementary Table 1. ICD-9 and ICD-10 Codes for maternal and neonatal diseases.

| Disease | ICD-9 codes | ICD-10 codes |
|---|---|---|
| **Multiple Sclerosis** | 340 | G35 |
| **Maternal complications** | | |
| Pre-gestational hypertension | 401-405, 642A-C, 642H +checkbox[a] | I10-I15, O10 and O11 +checkbox[a] |
| Preeclampsia and eclampsia | 642E-G | 014 and O15 |
| Pregestational diabetes | 250, 648A | E10 to E14, O24.0 to O24.3 |
| Gestational diabetes | 648W | 024.4 |
| **Maternal infection[b]** | 006-007X, 008W, 009-D, 084-086X, 099E-X, 110-136X, 321A, 321W, 370E-F, X, 372A-D, 380B, C, 381A, 382X, 420- 422X, 462-463, 466-B, 473-X, 483, 485-486, 490, 491B, 540A, X, 572A, 647C, E, W, X, 680A, 711G-X, 727A, 770A, 771C, E-W<br>Bacterial infection: 001-005X, 008A-F, 010-041X, 073, 076, 078D, J, 790H, 080-083X, 087-099D, 100-104, 245A, 254B, 320-X, 324-X, 360A, 373B, 375D, 376A, 382A-E, 383A-X, 390-392X, 421A, 461-X, 475, 481-482X, 510-X, 511B, 513-B, 522E, H, 526E, 527D, 528A, D, 540B, 562-B, 566, 567-C, 569F, 575A, 590-X, 597A, 595-D, X, 597W, 599A, 611A, 614-F, W-X, 615A, X, 616-X, 634A, 635A, 636A, 637A, 638A, 639A, 646F, G, 647A, B, D, 658E, 659D, 670, 675-B, W-X, 681-686X, 711A, E, 728A, 729E, 730-D, X, 771D, 996G, 998F, 999D<br>Viral infection: 008H-M, 045-066, 070-072X, 074-075, 077-078H, 078W-079X, 279K, 321B-H, 323A, 323C-D, 460, 464-465X, 480-X, 487-W, 647F, G, 711F, 771A, B, 790W | A06-07.9, A08.5, A09, A59-59.9, A63, A63.8-64, B35-49, B50 -89, B99, G02.1-02.8, G04, G04.9, G05.2, H10.0, H10.3-10.9, H16.2-16.3, H16.9, H32, H60, H60.3, H65.0-65.1, H66.9, I30.0-30.9, I33.0-33.9, I40.0, J02[c], J02.8-02.9[c], J03[c], J03.8-03.9[c], J16, J16.8, J18-18.9, J20, J20.8-21, J21.8-21.9, J22, J32-32.9[c], J35.0, J37-37.1[c], J40-42, K35, K35.9, K75.0, L30.3, M46.5, M65.1, M71.1, O98.3, O98.6-98.9, P23.8-23.9, P37.1-39.9, Z22.4, Z22.8-22.9, A00-05.9, A15-17.9, A20-28.9, A30 -58, A65 -79.9, B95-96.8, E06.0, E32.1, G00-00.9, G01, G04.2, G05.0, G06-06.2[c], G07, H00.0, H01.0, H04.3, H05.0, H44.0, H60.0-60.1, H66.0-66.4, H70.0-70.9, I00-02.9, J01-01.9[c], J02.0, J03.0, J13-15.9, J16.0, J20.0-20.2, J34.0, J36[c], J39.0-39.1, J85.1-85.3, J86-86.9[c], K04.6-04.7, K05.2, K11.3, K12.2, K14.0, K35.1, K57-57.9, K61-61.4, K63.0, K65.0a, K81.0, K85, L00 -08.9, M00-00.9, M46.3[c], M60.0[c], M86-86.9[c], N10-12[c], N13.6[c], N15.1, N15.9, N30-30.3[c], N30.8-30.9[c], N34-34.1[c], N39.0[c], N61, N70-76.8[c], N98.0, |

| | | |
|---|---|---|
| | | O07.0, O07.5, O08.0, O23-23.9, O41.1, O75.3, O85-86.8[c], O91-91.1, O98.0-98.2, P23.1-23.6, P36, P37.0, T80.2, T81.4, T82.6-82.7, T83.5-83.6, T84.5-84.7, T85.7, T88.0, Z22.0-22.3, A08-08.4, A60-60.9, A63.0, A80-89, A90-99, B00-06.0, B06.8-09, B15-19.9, B20-24, B25-34, B97-97.8, G02.0, G05.1, J00, J04-06.9[c], J10-11.8, J12-12.9, J20.3-20.7, J21.0, O35.3, O98.4-98.5, P23.0, P35, Z21, Z22.5-22.6 |
| **Neonatal infections**[b] | 771 | P35 to P39 |

a. Pre-gestational hypertension is also recorded in a checkbox in the prenatal record at first prenatal visit. b Diagnoses of maternal infections, neonatal infections, and asphyxia-related neonatal complications are from Medical Birth Register or the Patient Register (in-patient hospital care, admission date at 0-27 days of age). c. The additional codes, B95-97.8, denotes the infecting organism. Bacteria= B95-96.8, Virus= B97- 97.8.

Supplementary Table 2. The distribution of maternal characteristics across BMI categories.

| Maternal characteristics | Early-pregnancy BMI | | | | |
|---|---|---|---|---|---|
| | **Underweight** | **Normal weight** | **Overweight** | **Obese** | **Missing** |
| **Total, n (%)** | 34076 (2.2) | 829967 (54.4) | 307748 (20.2) | 121014 (7.9) | 232058 (15.2) |
| **Maternal age, y** | | | | | |
| Mean (SD) | 27.2 (5.1) | 29.3 (5.0) | 29.8 (5.1) | 29.7 (5.2) | 29.6 (5.3) |
| ≤19 | 1737 (5.1) | 16481 (2.0) | 4597 (1.5) | 1653 (1.4) | 5209 (2.2) |
| 20-24 | 9197 (27) | 126882 (15.3) | 44001 (14.3) | 18725 (15.5) | 35306 (15.2) |
| 25-29 | 12250 (35.9) | 291028 (35.1) | 101882 (33.1) | 40087 (33.1) | 74677 (32.2) |
| 30-34 | 8021 (23.5) | 270312 (32.6) | 100195 (32.6) | 37915 (31.3) | 74756 (32.2) |
| ≥35 | 2871 (8.4) | 125264 (15.1) | 57073 (18.5) | 22634 (18.7) | 42110 (18.1) |
| **Region of birth** | | | | | |
| Nordic | 26207 (76.9) | 718039 (86.5) | 259425 (84.3) | 103058 (85.2) | 188316 (81.2) |
| Non-nordic | 7848 (23) | 111565 (13.4) | 48148 (15.6) | 17904 (14.8) | 43520 (18.8) |
| Missing | 21 (0.1) | 363 (0.0) | 175 (0.1) | 52 (0.0) | 222 (0.1) |
| **Education, y** | | | | | |
| >12 | 14856 (43.6) | 437426 (52.7) | 137374 (44.6) | 43026 (35.6) | 112832 (48.6) |
| ≤12 | 19051 (55.9) | 390516 (47.1) | 169443 (55.1) | 77549 (64.1) | 117766 (50.7) |
| Missing | 169 (0.5) | 2025 (0.2) | 931 (0.3) | 439 (0.4) | 1460 (0.6) |
| **Mother cohabits with partner** | | | | | |
| Yes | 30202 (88.6) | 766653 (92.4) | 285224 (92.7) | 111187 (91.9) | 157108 (67.7) |
| No | 2678 (7.9) | 39273 (4.7) | 14893 (4.8) | 7042 (5.8) | 10617 (4.6) |
| Missing | 1196 (3.5) | 24041 (2.9) | 7631 (2.5) | 2785 (2.3) | 64333 (27.7) |
| **Parity** | | | | | |
| Nulliparous | 17323 (50.8) | 375142 (45.2) | 117051 (38.0) | 41908 (34.6) | 108350 (58.5) |
| Multiparous | 16753 (49.2) | 454825 (54.8) | 190697 (62.0) | 79106 (65.4) | 76949 (41.5) |
| **Maternal height, cm** | | | | | |
| Mean (SD) | 166.05 (6.52) | 166.59 (6.18) | 165.89 (6.29) | 165.55 (6.38) | 166.3 (6.3) |
| ≤159cm | 4977 (14.6) | 96639 (11.6) | 42905 (13.9) | 18673 (15.4) | 23215 (10.0) |
| 160-164 | 8414 (24.7) | 206853 (24.9) | 82484 (26.8) | 33420 (27.6) | 45086 (19.4) |
| 165-169 | 10065 (29.5) | 248823 (30.0) | 91186 (29.6) | 35306 (29.2) | 53290 (23.0) |
| ≥170 | 10620 (31.2) | 277652 (33.5) | 91173 (29.6) | 33615 (27.8) | 57229 (24.7) |
| **Maternal MS** | | | | | |
| No | 34060 (100) | 829366 (99.9) | 307549 (99.9) | 120906 (99.9) | 231914 (99.9) |
| Yes | 16 (0.0) | 600 (0.1) | 198 (0.1) | 108 (0.1) | 133 (0.1) |
| Missing | | <5 (0.0) | <5 (0.0) | | 11 (0.0) |
| **Smoking** | | | | | |
| No | 27179 (79.8) | 713311 (85.9) | 259544 (84.3) | 98199 (81.1) | 147154 (63.4) |
| Yes | 6304 (18.5) | 104219 (12.6) | 43512 (14.1) | 20957 (17.3) | 28016 (12.1) |
| Missing | 593 (1.7) | 12437 (1.5) | 4692 (1.5) | 1858 (1.5) | 56888 (24.5) |

Supplementary Table 3. Early-pregnancy body mass index and rates of multiple sclerosis in term appropriate-for-gestational-age births uncomplicated by maternal diabetes, hypertension, or preeclampsia

| **Maternal BMI** | **Number of MS cases** | **Person-years** | **Rates per 10000 person-years** | **Adjusted HR (95%CI)** |
|---|---|---|---|---|
| Underweight | 37 | 986255 | 0.37 | 0.86 (0.61-1.22) |
| Normal weight | 961 | 24941158 | 0.39 | Reference |
| Overweight | 381 | 8929200 | 0.43 | 1.16 (1.02-1.32) |
| Obesity | 160 | 3478006 | 0.46 | 1.33 (1.10-1.60) |

a. Adjusted for maternal age, parity, mother's country of origin, education level, cohabitation with partner, height, smoking status, maternal MS, and year of delivery.

Supplementary Table 4. Early-pregnancy body mass index and rates of multiple sclerosis in the overall cohort after multiple imputation.

| **Maternal BMI** | **Number of offspring** | **Number of MS cases** | **Person-years** | **Rates per 10000 person-years** | **Adjusted HR (95%CI)** |
|---|---|---|---|---|---|
| Underweight | 38775 | 37 | 986255 | 0.37 | 0.86 (0.61-1.22) |
| Normal weight | 985631 | 961 | 24941158 | 0.39 | Reference |
| Overweight | 358417 | 381 | 8929200 | 0.43 | 1.16 (1.02-1.32) |
| Obesity | 142039 | 160 | 3478006 | 0.46 | 1.33 (1.10-1.60) |

a. Adjusted for maternal age, parity, mother's country of origin, education level, cohabitation with partner, height, smoking status, maternal MS, and year of delivery.

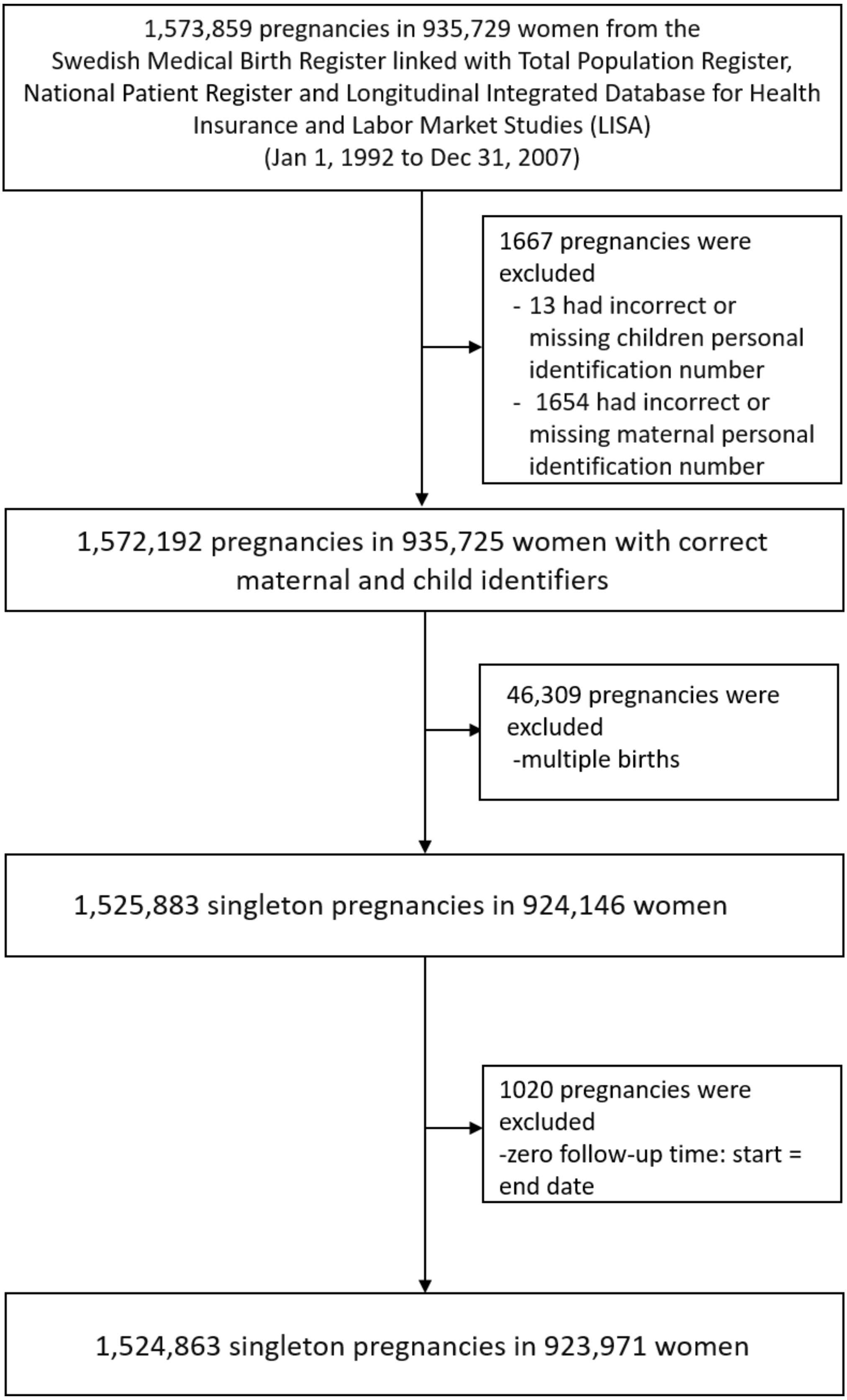


Supplementary Figure 1. Flowchart of inclusion criteria and final study population

| Maternal BMI | MS cases in offspring | Person-years | Rate per 10,000 person-years | Crude HR (95% CI) | Adjusted HR[a] (95% CI) |
|---|---|---|---|---|---|
| **Overall** | | | | | |
| Underweight | 21 | 625127 | 0.34 | 0.86 (0.56-1.33) | 0.81 (0.52-1.26) |
| Normal weight | 593 | 15846066 | 0.37 | Reference | Reference |
| Overweight | 216 | 5376402 | 0.40 | 1.13 (0.97-1.33) | 1.14 (0.97-1.34) |
| Obese | 87 | 1835710 | 0.47 | 1.42 (1.14-1.78) | 1.43 (1.13-1.82) |
| **POMS** | | | | | |
| Underweight | 1 | 426765 | 0.02 | 0.29 (0.04-2.06) | 0.29 (0.04-2.02) |
| Normal weight | 90 | 10994853 | 0.08 | Reference | Reference |
| Overweight | 31 | 3807580 | 0.08 | 0.99 (0.66-1.49) | 1.01 (0.67-1.53) |
| Obese | 17 | 1329413 | 0.13 | 1.56 (0.93-2.62) | 1.32 (0.74-2.35) |
| **AOMS** | | | | | |
| Underweight | 20 | 198363 | 1.01 | 0.96 (0.61-1.50) | 0.89 (0.56-1.41) |
| Normal weight | 503 | 4851015 | 1.04 | Reference | Reference |
| Overweight | 183 | 1568778 | 1.17 | 1.15 (0.97-1.36) | 1.16 (0.98-1.39) |
| Obese | 70 | 506262 | 1.38 | 1.39 (1.09-1.79) | 1.46 (1.13-1.90) |

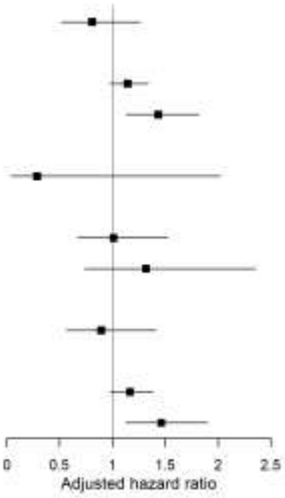


Supplementary Figure 2. Early-pregnancy body mass index and rates of multiple sclerosis in term appropriate-for-gestational-age births uncomplicated by maternal diabetes, hypertension, or preeclampsia

a. Adjusted for maternal age, parity, mother's country of origin, education level, cohabitation with partner, height, smoking status, maternal MS, and year of delivery.

### Panel A

| Maternal BMI | MS cases in offspring | Person-years | Rate per 10,000 person-years | Crude HR (95% CI) | Adjusted HR[a] (95% CI) |
|---|---|---|---|---|---|
| **Overall** | | | | | |
| Underweight | 13 | 434953 | 0.30 | 0.79 (0.45-1.38) | 0.64 (0.36-1.15) |
| Normal weight | 341 | 9364576 | 0.36 | Reference | Reference |
| Overweight | 103 | 2879082 | 0.36 | 1.04 (0.83-1.29) | 1.07 (0.86-1.34) |
| Obese | 42 | 1012280 | 0.41 | 1.28 (0.93-1.77) | 1.30 (0.93-1.81) |
| **POMS** | | | | | |
| Underweight | <5 | 300074 | 0.03 | 0.42 (0.06-3.06) | 0.37 (0.05-2.66) |
| Normal weight | 52 | 6553694 | 0.08 | Reference | Reference |
| Overweight | 13 | 2056251 | 0.06 | 0.79 (0.43-1.46) | 0.80 (0.43-1.49) |
| Obese | 8 | 739410 | 0.11 | 1.36 (0.64-2.85) | 1.20 (0.53-2.73) |
| **AOMS** | | | | | |
| Underweight | 12 | 134879 | 0.89 | 0.85 (0.48-1.52) | 0.69 (0.38-1.26) |
| Normal weight | 289 | 2810752 | 1.03 | Reference | Reference |
| Overweight | 90 | 822814 | 1.09 | 1.09 (0.86-1.38) | 1.12 (0.88-1.43) |
| Obese | 34 | 272860 | 1.25 | 1.27 (0.89-1.81) | 1.31 (0.91-1.89) |

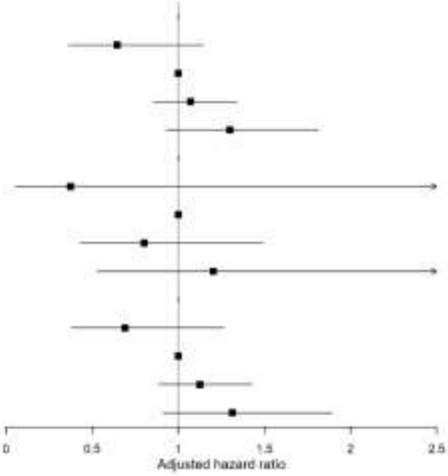


### Panel B

| Maternal BMI | MS cases in offspring | Person-years | Rate per 10,000 person-years | Crude HR (95% CI) | Adjusted HR[a] (95% CI) |
|---|---|---|---|---|---|
| **Overall** | | | | | |
| Underweight | 19 | 432205 | 0.44 | 1.12 (0.71-1.77) | 1.15 (0.72-1.82) |
| Normal weight | 435 | 11591431 | 0.38 | Reference | Reference |
| Overweight | 217 | 4758431 | 0.46 | 1.28 (1.09-1.51) | 1.29 (1.08-1.52) |
| Obese | 90 | 1930304 | 0.47 | 1.40 (1.11-1.75) | 1.40 (1.10-1.79) |
| **POMS** | | | | | |
| Underweight | <5 | 291148 | 0.07 | 0.87 (0.21-3.56) | 0.85 (0.21-3.49) |
| Normal weight | 63 | 7969950 | 0.08 | Reference | Reference |
| Overweight | 31 | 3344068 | 0.09 | 1.17 (0.76-1.80) | 1.16 (0.75-1.79) |
| Obese | 19 | 1389419 | 0.14 | 1.73 (1.04-2.89) | 1.49 (0.86-2.60) |
| **AOMS** | | | | | |
| Underweight | 17 | 141057 | 1.21 | 1.16 (0.71-1.88) | 1.20 (0.74-1.95) |
| Normal weight | 371 | 3621390 | 1.02 | Reference | Reference |
| Overweight | 184 | 1414310 | 1.30 | 1.29 (1.08-1.54) | 1.31 (1.09-1.58) |
| Obese | 71 | 540861 | 1.31 | 1.33 (1.03-1.72) | 1.39 (1.06-1.82) |

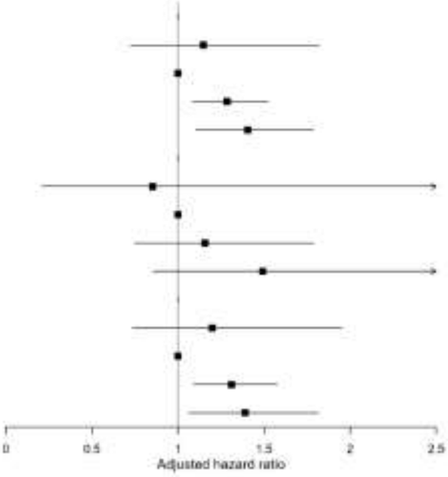


Supplementary Figure 3. Early-pregnancy body mass index and rates of multiple sclerosis in offspring among nulliparous (Panel A) and multiparous women (Panel B).

a. Adjusted for maternal age, parity, mother's country of origin, education level, cohabitation with partner, height, smoking status, maternal MS, and year of delivery.

**Panel A**

| Maternal BMI | MS cases in offspring | Person-years | Rate per 10,000 person-years | Crude HR (95% CI) | Adjusted HR[a] (95% CI) |
|---|---|---|---|---|---|
| **Overall** | | | | | |
| Underweight | 12 | 440228 | 0.27 | 1.21 (0.68-2.16) | 1.02 (0.56-1.87) |
| Normal weight | 233 | 10763637 | 0.22 | Reference | Reference |
| Overweight | 101 | 3926405 | 0.26 | 1.25 (0.99-1.58) | 1.26 (0.99-1.60) |
| Obese | 34 | 1515353 | 0.22 | 1.16 (0.81-1.67) | 1.14 (0.77-1.68) |
| **POMS** | | | | | |
| Underweight | <5 | 296727 | 0.03 | 0.71 (0.10-5.21) | 0.63 (0.09-4.45) |
| Normal weight | 35 | 7455813 | 0.05 | Reference | Reference |
| Overweight | 13 | 2774458 | 0.05 | 1.00 (0.53-1.88) | 1.01 (0.53-1.95) |
| Obese | 8 | 1096484 | 0.07 | 1.55 (0.72-3.34) | 1.21 (0.50-2.93) |
| **AOMS** | | | | | |
| Underweight | 11 | 140501 | 0.78 | 1.30 (0.71-2.38) | 1.10 (0.58-2.08) |
| Normal weight | 197 | 3307700 | 0.60 | Reference | Reference |
| Overweight | 68 | 1151932 | 0.76 | 1.31 (1.02-1.68) | 1.32 (1.02-1.71) |
| Obese | 26 | 418851 | 0.62 | 1.09 (0.72-1.63) | 1.13 (0.73-1.74) |

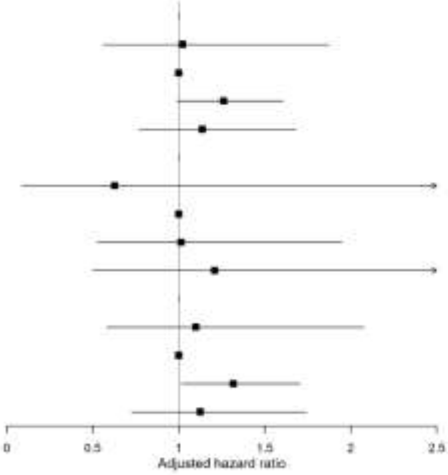


**Panel B**

| Maternal BMI | MS cases in offspring | Person-years | Rate per 10,000 person-years | Crude HR (95% CI) | Adjusted HR[a] (95% CI) |
|---|---|---|---|---|---|
| **Overall** | | | | | |
| Underweight | 20 | 426930 | 0.47 | 0.85 (0.54-1.32) | 0.83 (0.53-1.30) |
| Normal weight | 543 | 10192370 | 0.53 | Reference | Reference |
| Overweight | 219 | 3711107 | 0.59 | 1.17 (1.00-1.37) | 1.17 (1.00-1.38) |
| Obese | 98 | 1427202 | 0.69 | 1.44 (1.16-1.79) | 1.45 (1.15-1.81) |
| **POMS** | | | | | |
| Underweight | <5 | 291495 | 0.07 | 0.61 (0.15-2.48) | 0.58 (0.14-2.40) |
| Normal weight | 80 | 7067831 | 0.11 | Reference | Reference |
| Overweight | 31 | 2625861 | 0.12 | 1.04 (0.69-1.58) | 1.02 (0.67-1.54) |
| Obese | 19 | 1032326 | 0.18 | 1.82 (0.98-2.68) | 1.45 (0.85-2.47) |
| **AOMS** | | | | | |
| Underweight | 18 | 135435 | 1.33 | 0.88 (0.55-1.42) | 0.87 (0.54-1.40) |
| Normal weight | 463 | 3124441 | 1.48 | Reference | Reference |
| Overweight | 186 | 1085192 | 1.71 | 1.18 (0.99-1.40) | 1.20 (1.00-1.43) |
| Obese | 79 | 394859 | 2.00 | 1.40 (1.11-1.78) | 1.44 (1.12-1.85) |

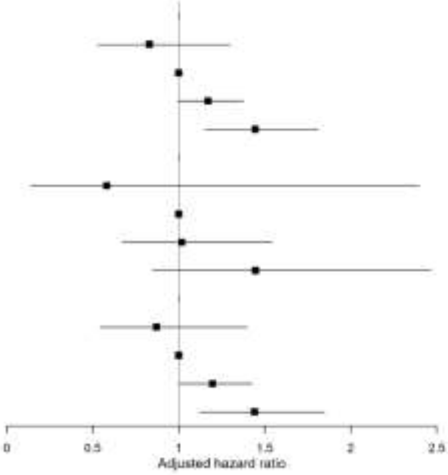


**Supplementary Figure 4**. Early-pregnancy body mass index and rates of multiple sclerosis in male (Panel A) versus female (Panel B) offspring

a. Adjusted for maternal age, parity, mother's country of origin, education level, cohabitation with partner, height, smoking status, maternal MS, and year of delivery.
